# Predictors of misconceptions, knowledge, attitudes, and practices of COVID-19 pandemic among a sample of Saudi population and its impact: a cross-sectional study

**DOI:** 10.1101/2020.05.22.20110627

**Authors:** Mukhtiar Baig, Tahir Jameel, Sami H Alzahrani, Ahmad A Mirza, Zohair J Gazzaz, Tauseef Ahmad, Fizzah Baig, Saleh H Almurashi

## Abstract

**Objectives:** To explore the predictors of misconceptions, knowledge, attitudes, and practices of the COVID-19 pandemic among a sample of the Saudi population and its impact.

**Design:** An online cross-sectional study.

**Setting:** Jeddah, Saudi Arabia.

**Participants:** A total of 2006 participants [953(47.5%) females, and 1053(52.5%) males], and more than 18 years of age were included in the study.

**Data collection and analysis:** This cross-sectional survey was conducted at the Faculty of Medicine, Rabigh, King Abdulaziz University (KAU). The participants were approached by social media (SM). An online questionnaire was administered, and the data were analyzed on SPSS-26.

**Results:** The SM was the leading source of information 889(43.9%). Two-thirds of the participants, 1250(66.9%) had various misconceptions, and about one-third of the study participants 637(31.7%) had disturbed social, mental, and psychological wellbeing, and many participants became more religious. Two-thirds of the study participants, 1292(68.1%) had good knowledge score. The attitude was highly positive in 1867(93.1%) participants’, and the practice score was adequate in 1939(97.7%). The participants’ educational status was the predictor of good knowledge. Male gender and divorced ones were the predictor of poor practice scores and age 51-60 years, private jobholders, and students were the predictors of the good practice scores. The Saudi nationality was the predictor of participants’ positive attitude, while the male gender and divorced ones were predictors of a negative attitude. The male gender and higher education status were the predictors of good concepts, while the older age and job (own business) were the predictors of misconceptions.

**Conclusion:** Our study observed several predictors of misconceptions, knowledge, attitudes, and practices among the Saudi population. Our participants had a good realization of the impact of this pandemic.

**Strengths and limitations:**

- Ours is the first study involving the general population of Saudi Arabia at a time when COVID-19 has tremendously affected the society.
- The use of a validated questionnaire has strengthened the study results.
- The sample size is adequate and representing different segments of society.
- One of the limitations is the convenience sampling method.
- Besides, the study used an online questionnaire, so we could not reach the section of society that is not using the internet.

## INTRODUCTION

In recent years, Coronaviruses have become a major health hazard throughout the world by causing considerable morbidity in the human. In a short period, novel Coronavirus Disease 2019 (COVID-19) has transformed from endemic to a pandemic. This infection has been transmitted to 213 countries and territories in the world and infected 5,168,433 people with 335,936 mortalities (22 May 2020, 10:20 PM).^1^ It has dilapidated everything from the economy to social life worldwide. Initially, the COVID-19 pandemic was symbolized by the ignorance, mayhem, repudiation, and fright. However, it swarmed with an unbelievably rapid pace, infecting thousands of people worldwide. Most of the countries had to lock down their cities, then people noticed it seriously and started taking precautionary measures.^2^

Saudi Arabia has witnessed a variety of disturbing epidemics of Coronaviruses in recent few years such as SARS-CoV (Severe Respiratory Distress Syndrome-Corona Virus) in 2002 and MERS-CoV (Middle Eastern Respiratory Distress Syndrome-Corona Virus) in 2012.^3,4^ COVID-19, is a newer member of this family, has become a deadly pandemic within a very short time. Saudi Arabia is also severely hit by this deadly virus. Cases have been recorded in almost all the regions with pocketing of cases in multiple cities.

Authorities took drastic preventive and curative measures, including the phase-by-phase lockdown and imposition of curfew during the evening in almost all the major cities.^5^

Even though these measures are proving less effective, curfew has been extended to almost 24 hours with a brief relief for buying essential commodities. Till today (22 May 2020, 10:20 PM) the number of COVID-19 positive cases have increased to 67,719 with 364 deaths due to this disease.^1^

The control of communicable diseases depends to a large extent on the knowledge, attitudes, practices, and behavior of the local population.^6^ The strict observance of precautionary measures to avoid its spread to the masses is key to control the disease. Mass media is the major source of spreading different myths regarding prevention and resistance against the disease. The practicing behavior and knowledge of the general public carry the most important bearing in the war against COVID-19. Assessing the gap in general public misconceptions, knowledge, attitudes, and practice behaviors are always advantageous for developing future policies towards such pandemic situations.^7^ People have been overburdened by the influx of information from different resources, especially by the SM; thus, they are confused and anxious to find accurate knowledge. ^8, 9^

The present survey explored the predictors of misconceptions, knowledge, attitudes, and practices of the COVID-19 pandemic among a sample of the Saudi population. We also assessed their approach towards the overall impact of this problem. Our results would help to update the awareness campaigns accordingly.

## METHODS

The present cross-sectional questionnaire-based survey was conducted at the Faculty of Medicine, Rabigh, KAU, Jeddah, after taking ethical approval from the Unit of Biomedical Ethics of the University (Ref No. 187-20). The Rao soft sample size calculator calculated the sample size by considering the margin of error 5%, confidence level 95%, population size 3000000, the required sample size was 385. The convenience sample technique was used, and participants were approached by SM (Facebook, Whatsapp, Twitter, and others), and an online questionnaire was administered, and no monetary benefit was offered to any participants. At the beginning of the online questionnaire, a brief description of the research was given, and a request for their participation. Participants agreed to fill the online questionnaire was considered their consent for participation in the survey. The questionnaire was constructed with the help of the WHO myth-buster document and a published study.^10, 11^

The questionnaire was translated, and back-translated (English-Arabic) by two bilingual experts’ and the questionnaire was modified according to their suggestions. Two senior faculty members validated the questionnaire. A pilot study on 35 people from the general public was done to find out the convenience and comprehension of the questionnaire and its Cronbach’s alpha was .81. This questionnaire was converted to a google document containing English and Arabic translation. We kept both languages together so that everybody can fill the questionnaire according to its convenience. This questionnaire was converted to a google document, and the link was sent via SM like WhatsApp’s, Twitter, Facebook, and others to all the participants.

People having age less than 18 years and residing outside Jeddah were not included in the study. Our questionnaire had several parts; the first part comprised of the demographic questions, and then there were 14 knowledge questions, 6 attitude, 4 practice, 19 misconceptions, and 6 impact questions. Knowledge, attitude, misconceptions, and impact of the outbreak were true/ false/not sure types, while practice questions were yes/no/sometimes types. “One score was awarded for true, and zero for false and not sure and an individual score less than 50% (1-7 score) was considered poor, while individual score 51% - 75% (8-10 score) was considered moderate, and 76-100% (11-14 score) was considered good. For attitude, marking ranged from −4 to +4 (true answer +1 and false and not sure −1), and individuals score in plus indicated a positive attitude while in minus indicated a negative attitude. The practice score ranged from 0-12 (yes=2 points, sometimes=1 and No= 0) and =>6 score was considered adequate and <6 was considered not adequate.” The misconception questions score ranged from 1-19 [correct (true) answer = 1 score, wrong (false) answer = 0 score, not sure= 0 score] and individual who received ≤ 50% score (1-9 score) were taken as poor and individual with > 50% score (10 – 19 score) was considered high score and it indicated good concepts.

### Statistical analysis

The collected data was analyzed on SPSS-26. The frequency and percentages were computed for different variables. A Chi-squared test was used to explore the comparison between different variables. Association of knowledge, attitudes, practices, and misconceptions scores with different variables was calculated by binary logistic and multiple logistic regression analysis. The p-values were considered significant if it was <0.05.

## RESULTS

A total of 2117 participants filled the questionnaire, but after removing incomplete responses, 2006 participants [953(47.5%) females, and 1053(52.5%)] were included in the study. According to job nature 808(40.3%), 298(14.9%), 65(3.2%), 294(14.7%), and 541(27%) were government servants, private jobs, own business, housewives, and students, respectively. Among our participants, the sources for seeking information of COVID-19 were SM 889(43.9%), govt services 653(32.1%), TV 320(15.7%), newspaper 44(2.2%), and other 130(6.3%) (Figure 1).

**Figure 1.**
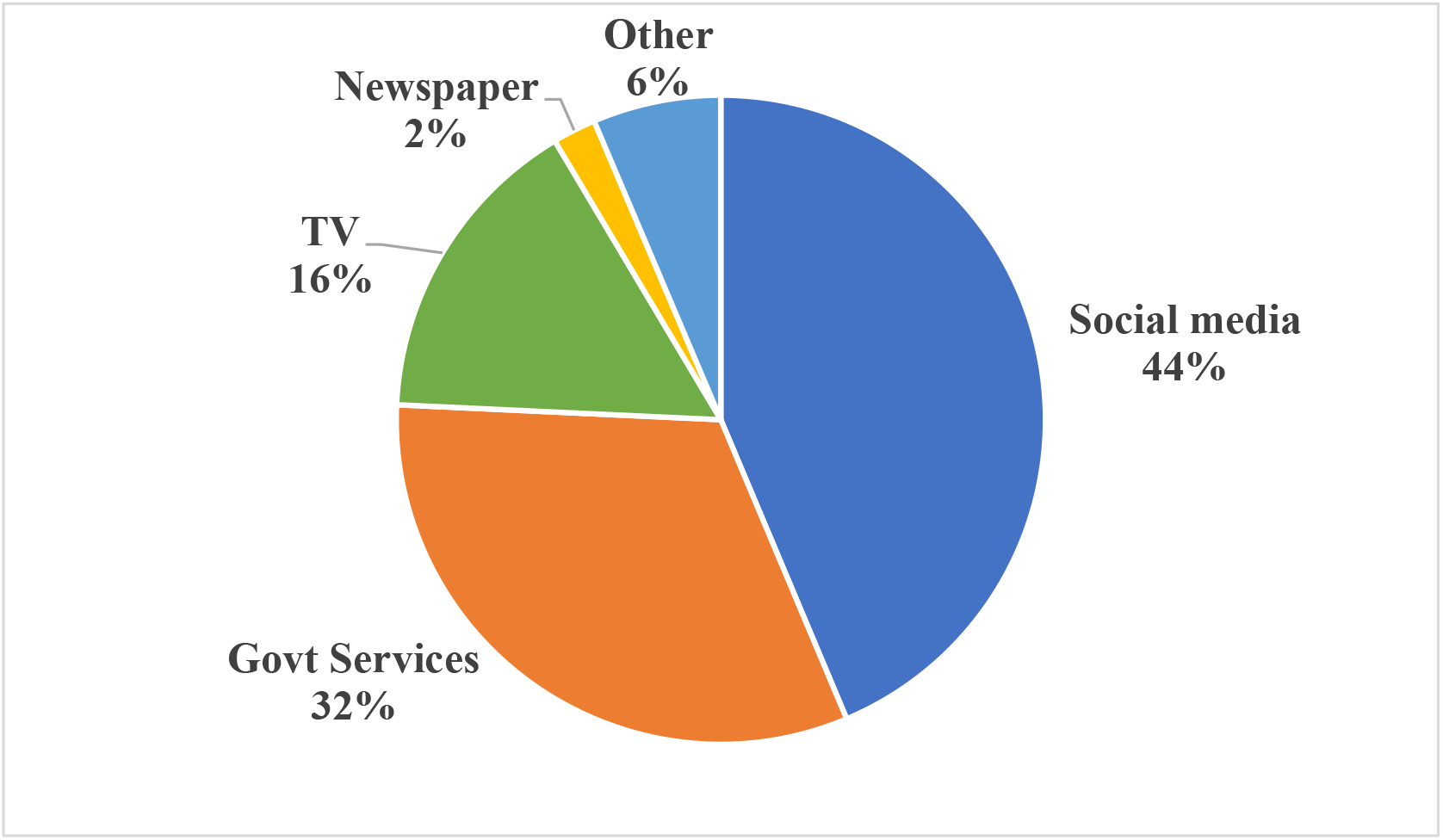
Source of information for study participants.

The participants’ responses regarding statements towards knowledge, attitudes, practices, misconceptions, and impact of COVID-19 pandemic are shown in Tables S1 and S2.

The few common misconceptions found were “females are more vulnerable to develop this infection 1067(56.2%), rinsing the nose with saline and sipping water every 15 minutes protects against coronavirus [899(47.4%);825(43.5%), respectively], flu and pneumonia vaccines protect against this virus 966(50.9%)”. About half of the respondents 925(46.3%) were terrified of COVID-19, and about one-third 637(31.7%) of the study participants had disturbed social, mental, and psychological wellbeing, and many participants became more religious (Table S2). Two-third of the study participants 1292(68.1%) had good knowledge of COVID19, 505(26.6%) had moderate knowledge. The attitude of the majority of the participants [1867(93.1%)] was highly positive, and the practice score of almost all the participants was adequate [1939(97.7%)]. The majority of the study participants (66.5%) had misconception [score <50 (poor)] while there were one-third participants who had good concepts [score >50 (good)] (33.5%) (Table 1).

**Table 1.**
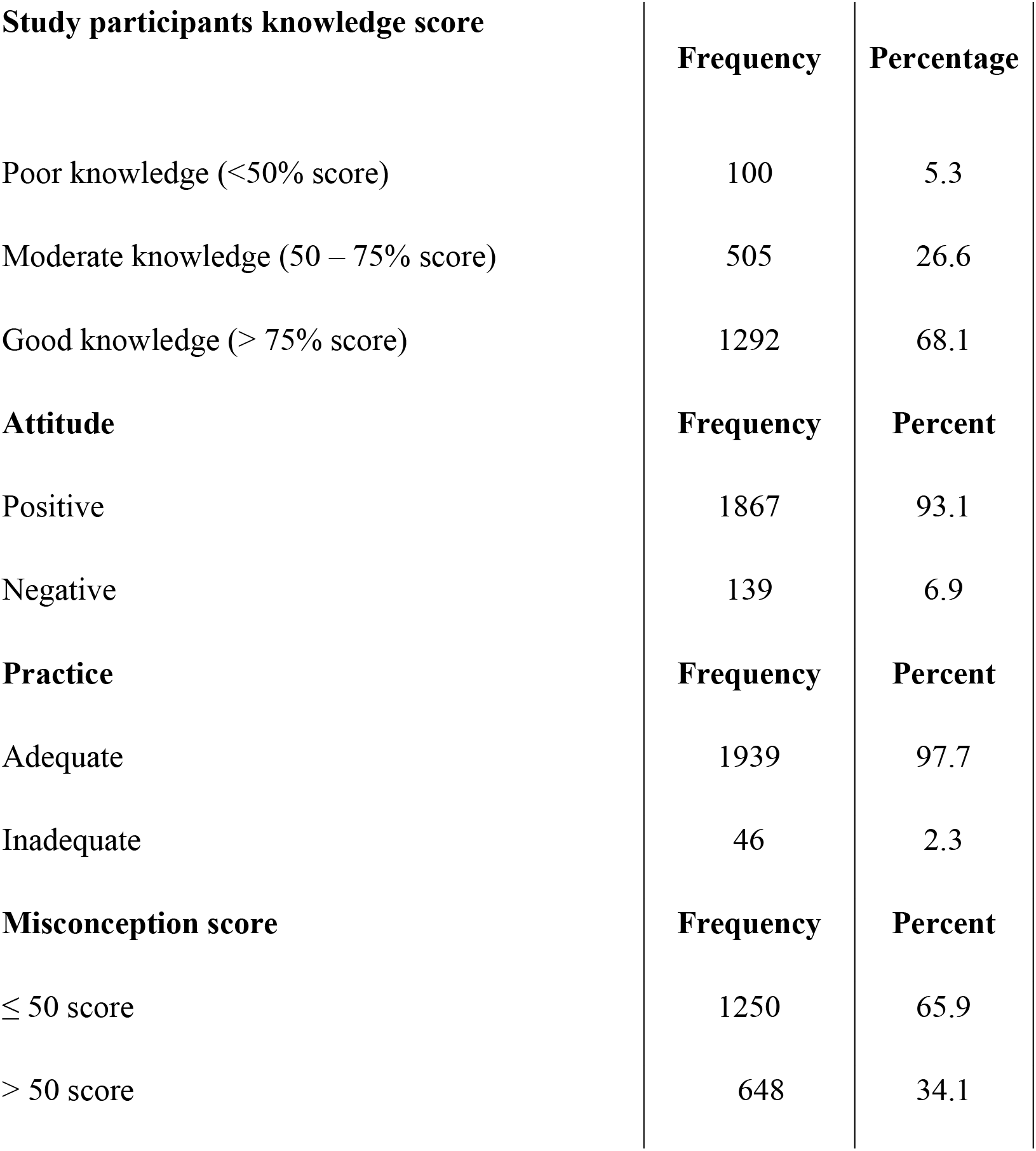
Study participants knowledge, attitude, practice and misconceptions scores.

Positive score indicates positive attitude, Negative or zero score indicates negative attitude Correct (true) answer = 1 score, Wrong (false) answer = 0 score, not sure= 0 score ≤ 50% score = Poor (1 – 9 score), > 50% score= good (10 – 19 score), High score indication good concept

More participants’ in the younger age group had a good knowledge score (p=.037), and more educated people had better knowledge (p<.001). A significant difference of attitude was observed according to gender (females were more positive), nationality (Saudis were more positive), education (except masters all were more positive), and marital status (married were more positive). More participants’ in the age group 40-50 yrs., 51-60 yrs., and >60 yrs showed good practice (p=.041), and females also showed good practice as compared to the males (p<.001). More participants’ in the age group 18-28 yrs., and 29-39 yrs., showed good concepts compared to other age groups (p<.001), and males also showed good concepts as compared to females (p<.001). Highly educated people (college, master, Ph.D.) showed good concepts compared to primary school and high school people (p<.001). Students, private jobs, and govt job people had good concepts compared to housewives and own business owners (p<.001). According to marital status, unmarried had better concepts compared to married and divorced (p<.001) (Table 2).

**Table 2.** Comparison of knowledge, attitude, practice and misconception scores according to different variables.

| Variables |  | Knowledge |  |  | Attitude |  | Practice |  | Misconception |  |
| --- | --- | --- | --- | --- | --- | --- | --- | --- | --- | --- |
|  |  | Poor | Moderate | Good | Negative | Positive | Inadequate | Adequate | ≤ 50 score | > 50 score |
| Age | 18-28 | 37<br>(5.0%) | 162<br>(22.0%) | 536<br>(72.9%) | 55 (7.1%) | 716<br>(92.9%) | 22 (2.9%) | 743<br>(97.1%) | 403 (55.4%) | 324<br>(44.6%) |
|  | 29-39 | 29<br>(5.7%) | 142<br>(28.0%) | 337<br>(66.3%) | 44 (8.4%) | 480<br>(91.6%) | 17 (3.3%) | 499<br>(96.7%) | 316 (64.6%) | 173<br>(35.4%) |
|  | 40-50 | 23<br>(5.0%) | 138<br>(29.8%) | 302<br>(65.2%) | 33 (6.6%) | 466<br>(93.4%) | 7 (1.4%) | 486<br>(98.64%) | 370 (77.4%) | 108<br>(22.6%) |
|  | 51-60 | 10<br>(6.3%) | 52 (32.5%) | 98<br>(61.3%) | 6 (3.4%) | 169<br>(32.5%) | 0 (0.0%) | 174<br>(100%) | 132 (78.1%) | 37<br>(21.9%) |
|  | >60 | 1 (3.2%) | 11 (35.5%) | 19<br>(61.3%) | 1 (2.7%) | 36<br>(97.3%) | 0 (0.0%) | 37 (100%) | 29 (82.9%) | 6<br>(17.1%) |
| p-value | 0.037* |  |  |  | 0.183 |  | 0.041 |  | < 0.001* |  |
| Gender | Male | 60<br>(6.0%) | 269<br>(27.0%) | 668<br>(67.0%) | 89 (8.5%) | 957<br>(91.5%) | 41 (3.9%) | 1004<br>(96.1%) | 614 (61.6%) | 382<br>(38.4%) |
|  | Female | 40<br>(4.4%) | 236<br>(26.2%) | 624<br>(69.3%) | 50 (5.3%) | 901<br>(94.7%) | 5 (0.5%) | 928<br>(99.5%) | 630 (70.8%) | 363<br>(29.5%) |
| p-value | 0.259 |  |  |  | 0.004* |  | < 0.001* |  | < 0.001* |  |
| Nationality | Saudi | 79<br>(4.9%) | 429<br>(26.4%) | 1114<br>(68.7%) | 102<br>(6.0%) | 1608<br>(94.0%) | 43 (2.5%) | 1649<br>(97.5%) | 545 (33.8%) | 1614<br>(100%) |
|  | Non Saudi | 21<br>(7.6%) | 76 (27.6%) | 178<br>(64.7%) | 36<br>(12.5%) | 251<br>(87.5%) | 3 (1.1%) | 281<br>(98.9%) | 101 (36.7%) | 275<br>(100%) |
| p-value | 0.130 |  |  |  | < 0.001* |  | 0.125 |  | 0.339 |  |
| Education | Primary school | 2<br>(11.1%) | 7 (38.9%) | 9<br>(50.0%) | 1 (4.5%) | 21<br>(95.5%) | 0 (0%) | 22 (100%) | 18 (94.7%) | 1 (5.3%) |
|  | College | 58<br>(4.4%) | 318<br>(24.3%) | 932<br>(71.3%) | 86 (6.3%) | 1289<br>(93.7%) | 27 (2.0%) | 1334<br>(98.0%) | 811 (62.3%) | 491<br>(37.7%) |
|  | High school graduate | 19<br>(5.6%) | 132<br>(38.9%) | 188<br>(55.5%) | 16 (4.4%) | 344<br>(95.6%) | 8 (2.2%) | 348<br>(97.8%) | 281 (81.4%) | 64<br>(18.6%) |
|  | Master | 15<br>(8.9%) | 33 (19.6%) | 120<br>(71.4%) | 30<br>(17.0%) | 146<br>(83.0%) | 8 (4.6%) | 166<br>(95.4%) | 102 (61.8%) | 63<br>(38.2%) |
|  | Ph.D | 6 (9.4%) | 15 (23.4%) | 43<br>(67.2%) | 6 (9.1%) | 60<br>(90.9%) | 3 (4.6%) | 62<br>(95.4%) | 36 (59.0%) | 25<br>(41.0%) |
| p-value | < 0.001 |  |  |  | < 0.001* |  | 0.153 |  | < 0.001* |  |
| Job | Govt job | 40<br>(5.2%) | 215<br>(28.0%) | 513<br>(66.8%) | 52 (6.4%) | 756<br>(93.6%) | 23 (2.9%) | 779<br>(97.1%) | 520 (68.0%) | 245<br>(32.0%) |
|  | Private job | 16<br>(5.6%) | 76 (26.4%) | 196<br>(68.1%) | 23 (7.8%) | 273<br>(92.2%) | 4 (1.4%) | 291<br>(98.6%) | 173 (62.0%) | 106<br>(38.0%) |
|  | own business | 3 (5.8%) | 18 (34.6%) | 31<br>(59.6%) | 6 (9.5%) | 57<br>(90.5%) | 3 (4.7%) | 61<br>(95.3%) | 52 (82.5%) | 11<br>(17.5%) |
|  | housewife | 16<br>(5.9%) | 84 (31.2%) | 169<br>(62.8%) | 16 (5.5%) | 277<br>(94.5%) | 1 (0.4%) | 283<br>(99.6%) | 220 (80.0%) | 55<br>(20.0%) |
|  | student | 25<br>(4.8%) | 112<br>(21.5%) | 383<br>(73.7%) | 41 (7.6%) | 497<br>(92.4%) | 15 (2.8%) | 518<br>(97.2%) | 278 (54.7%) | 230<br>(45.3%) |
| p-value | 0.090 |  |  |  | 0.614 |  | 0.055 |  | < 0.001* |  |
| Marital Status | Married | 60<br>(5.4%) | 308<br>(27.8%) | 739<br>(66.8%) | 74 (6.3%) | 1098<br>(93.7%) | 19 (1.6%) | 1141<br>(98.4%) | 797 (71.9%) | 312<br>(28.1%) |
|  | Unmarried | 36<br>(4.9%) | 179<br>(24.1%) | 527<br>(71.0%) | 56 (7.3%) | 715<br>(92.7%) | 25 (3.3%) | 740<br>(96.7%) | 406 (55.7%) | 323<br>(44.3%) |
|  | Divorced | 4 (8.3%) | 18 (37.5%) | 26<br>(54.2%) | 9 (16.4%) | 46<br>(83.6%) | 2 (3.8%) | 50<br>(96.1%) | 40 (76.9%) | 12<br>(23.1%) |
| p-value | 0.084 |  |  |  | 0.015* |  | 0.052 |  | < 0.001* |  |

Multiple regressions analysis showed that the participants’ educational status was the predictor of good knowledge. Male gender and divorced ones were the predictor of poor practice scores and age 51-60 years, private jobholders, and students were the predictors of the good practice scores (Table 3).

**Table 3.** Multiple regression analysis showing predictors of knowledge and practice.

| Variables | Knowledge |  | Practice |  |
| --- | --- | --- | --- | --- |
|  | Model 1<sup>a</sup> |  |  |  |
|  | B | P-value | B | P-value |
| Age |  |  |  |  |
| 29-39 | -.087 | .605 | .056 | .707 |
| 40-50 | -.041 | .825 | .169 | .295 |
| 51-60 | -.271 | .232 | .462 | .020 |
| >60 | .091 | .813 | .609 | .061 |
| Gender |  |  |  |  |
| Male | -.197 | .051 | -1.115 | .000 |
| Nationality |  |  |  |  |
| Saudi | .118 | .402 | -.217 | .083 |
| Educational status |  |  |  |  |
| College | .852 | .067 | -.405 | .234 |
| High school graduate | .343 | .468 | -.340 | .328 |
| Master | .738 | .129 | -.612 | .092 |
| Ph.D. | 1.215 | .021 | -.240 | .554 |
| Job status |  |  |  |  |
| Private job | .028 | .852 | .413 | .002 |
| own business | -.356 | .210 | .275 | .242 |
| housewife | -.227 | .156 | .031 | .826 |
| student | .180 | .305 | .337 | .028 |
| Marital status |  |  |  |  |
| Unmarried | -.043 | .780 | -.179 | .188 |
| Divorced | -.559 | .052 | -.503 | .045 |

a=include age, gender, nationality, education, job, and marital status

Binary regression analysis revealed that the Saudi nationality was the predictor of participants’ positive attitude, while the male gender and divorced ones were predictors of a negative attitude. The male gender and higher education status were the predictors of good concepts, while the older age and job (own businesses) were the predictors of misconceptions. (Table 4).

**Table 4.** Binary logistic regression analysis Association of attitude and misconception scores with different variables.

| Variables | Attitude |  |  | Misconception |  |  |
| --- | --- | --- | --- | --- | --- | --- |
|  | B | P-value | OR | B | P-value | OR |
| 18-28 | Reference |  |  |  |  |  |
| 29-39 | -.253 | 0.447 | .777 | -.279 | .129 | .757 |
| 40-50 | .060 | 0.873 | 1.061 | -.811 | .000 | .445 |
| 51-60 | .925 | 0.089 | 2.522 | -.891 | .001 | .410 |
| >60 | 1.273 | 0.244 | 3.571 | -1.191 | .027 | .304 |
| Female | Reference |  |  |  |  |  |
| Male | -.688 | 0.002 | .503 | .272 | .017 | 1.312 |
| Non-Saudi | Reference |  |  |  |  |  |
| Saudi | .856 | 0.001 | 2.354 | -.285 | .073 | .752 |
| Primary school | Reference |  |  |  |  |  |
| College | -.171 | 0.870 | .843 | 2.096 | .044 | 8.135 |
| High school graduate | .130 | 0.903 | 1.139 | 1.209 | .250 | 3.350 |
| Master | -1.370 | 0.195 | .254 | 2.295 | .029 | 9.920 |
| Ph.D | -.623 | 0.582 | .536 | 2.462 | .022 | 11.724 |
| Govt job | Reference |  |  |  |  |  |
| Private job | .176 | 0.562 | 1.193 | -.017 | .920 | .983 |
| own business | -.436 | 0.374 | .647 | -1.120 | .006 | .326 |
| housewife | -.395 | 0.276 | .674 | -.317 | .117 | .729 |
| student | -.548 | 0.119 | .578 | .079 | .682 | 1.082 |
| Married | Reference |  |  |  |  |  |
| Unmarried | -.035 | 0.908 | .965 | .158 | .353 | 1.172 |
| Divorced | -1.248 | 0.002 | .287 | -.308 | .383 | .735 |

## DISCUSSION

The awareness and positive response of a society is the key to the successful handling of an emergency like COVID-19 pandemic. The spread of a pandemic infection creates a lot of anxiety and fear in society that needs to be assessed and treated.

### Source of information

Among our participants, the leading source of information of COVID-19 was SM, followed by govt websites, TV, newspaper, and others. Our results are similar to another study.^12^ The SM has become a substantial channel for news and information in the contemporary media milieu and millions of people in the world using it for updating knowledge. ^13^

The SM has extensive “health misinformation,” which is often described as information that contradicts existing evidence from medical specialists at the time.^14^ Therefore, its duty of information seeker to search medical information from reliable resources like WHO, CDC, and their MOH portals.

In addition to many positive aspects, the myths, gossips, and misinformation can quickly spread online, mainly via SM. People are more prone to disperse fake stories and fabricated information, and these fabricated stories circulate extensively and swiftly than factual narratives.^15^ It is worrying that the majority of our study participants, during this pandemic, were seeking information from SM. However, one-third of the study population were using government services for finding information. It is worth mentioning the MOH in SA is working very efficiently since day one, and its portal has updated information regarding COVID-19.

### Misconceptions

Several misconceptions were present among our participants. Two-thirds of the study participants (66.5%) had misconceptions. Scarce data are available about the misconceptions associated with the demographic variables. Our results revealed that the male gender and higher education status were the predictors of good concepts, while the older age and job (own businesses) were the predictors of misconceptions. Similar to our results, few other studies have reported misconceptions among their study participants.^16,17^ An Australian and multinational study reported that uncertainties and misconceptions about COVID-19 were widespread among the general public.^12,16^ Such misconceptions and misinformation could be the obstacles against taking appropriate precautionary measures and positive behavior change among the masses. The identification of accurate knowledge about a disease is as essential for risk-reduction behavior, and most interventions have concentrated on information propagation as the imperative phase in reducing disease risk.^18^

As the males are going outside for a job and finding more chances to discuss this issue with colleagues, thus their concepts regarding COVID-19 were clear. Educated people have more exposure and excess to knowledge; that’s why they had clear concepts. We found that the older age and people doing their businesses were the predictors of misconception, but we didn’t find any plausible explanation for this association. It is suggested that continuous efforts are needed to clarify the peoples’ misconceptions, and in some cases, by involving religious leaders, especially in Muslim countries. The most important thing is the sustained awareness campaigns by government and non-governmental organizations (NGOs) to bust these myths that are getting harmful to the societies.

### Impact

The current pandemic of COVID-19 has resulted in drastic effects on the economic horizon, the world over. There are predictions that the world economy and tourism would suffer a lot in the coming years. Millions of people have gone jobless, businesses are down, and almost every aspect of life is on a standstill.^19^ About half of the respondents, 925(46.3%) were terrified of COVID-19, and about one-third of the study participants 637(31.7%) had disturbed social, mental, and psychological wellbeing. The majority of the participants stated that this pandemic made them realized the importance of life, and one-third committed to becoming more religious. Most of our respondents stated that the overall impact of this pandemic would be on the country’s economic condition, followed by the healthcare system and the people’s financial status.

Interestingly, another study reported inclinations of the people towards religion during this pandemic issue. They were also embracing a healthy lifestyle and keeping them away from the mass gathering and praying at home instead of mosques.^20^ Gros et al. (2020) have also raised awareness and concerns among their study participants regarding the economic situation during this pandemic.^21^ A Chinese study reported similar impacts of this pandemic among its population.^22^ Another study emphasized for taking steps to tackle mental health problems like anxiety and depression during this COVID-19 pandemic. ^23^ People were scared and had mental distress, and these are explicable as health is a serious matter for the people. Nobody intends to have COVID-19 infection, which has a moderately high risk of mortality.^24^ Such results emphasize urging the people to adopt manners that reduce the infection risk and alleviate the psychological effects of the outbreak.^12, 25^ The participants’ reasons for good knowledge, attitudes, and practices could be that the survey was conducted at the advanced stage of the pandemic, and people’s awareness had increased. There is a need to develop a policy to treat the people who are suffering from mental illnesses because of the havoc caused by this virus.

### Knowledge

In our study, the Saudi population (95% had good to moderate knowledge). More participants in the younger age group and educated people had a good knowledge score. The participants’ educational status was the predictor of good knowledge. Our results are similar to several other studies. ^8,16,17^ An Australian study described the general public’s good knowledge in Australia, but they had knowledge gaps for a few questions^12^. Our results found no significant association between participants’ knowledge score and other variables, while in contrast to our results, few studies found a significant association between knowledge and demographic characteristics like age, gender, and occupation.^11,17^ The level of awareness was much higher among educated people, which is a common finding in other studies. ^26,27^ The knowledge of a person is transformed into attitude and later on reflected in practice. A survey from Arabic speaking Middle Eastern countries pointed out several gaps in public knowledge about COVID-19 and proposed health education for amending their knowledge^16^ and similar suggestions given by a Chinese study.^11^

### Attitudes

The majority of our study participants had a positive attitude (93.1%) and believed that this virus pandemic could be overcome by taking precautionary steps (93.6%). Ninety-five percent of the respondents agreed that infection is highly contagious, while 97.6% agreed that it’s our social responsibility to take safety measures in controlling the spread of this infection. More than half of the participants (56.1%) were optimistic that COVID-19 infection would be overcome soon. Malaysian and Chinese studies have also reported a very positive attitude among participants for similar questions.^8, 11^

The high-level positive attitude in our study may be attributed to an excellent campaign for the awareness of the Saudi population by the MOH. They are sending daily awareness messages on mobile phones in different languages and have launched a mobile application to identify the symptoms of the corona. Newspapers and television are also disseminating information regarding preventive measures. Such a good attitude among people was also attributed to the government’s efforts to mitigate the virus transmission by Chinese and Malaysian studies.^8, 11^

The Saudi nationality was the predictor of participants’ positive attitude, while the male gender and divorced ones were predictors of a negative attitude. One of the reasons could be a better living style and exposure to awareness campaigns on SM and local TV networks by Saudi nationals. Most of the expatriates belonged to the working class, and they were not highly educated. Hence, they had relatively insufficient knowledge regarding the disease as compared to Saudi nationals, which is also reflected in their attitude. Interestingly this finding has also been highlighted in other studies that females show more concern and positivity towards any infectious epidemic for their families and society.^28,29^

Education and marriage modify individual’s responses resulting in responsible attitude and overall positiveness, ^30^ so was observed in our study with some exceptions. One interesting finding, the divorced status was associated with a careless and rather negative attitude towards COVID-19. Nasser et al. (2020) mentioned similar results in their participants.^16^ The possible explanation may be that this group is a bit careless and indifferent towards even serious matters.

### Practices

Almost all the participants attained adequate scores regarding adopting protective measures (97.7%). Our study participants were maintaining social distancing by avoiding meeting with friends and relatives (89.9%), and not visiting crowded places (95.1%). Most of the participants were using soap frequently for handwashing (91%), using face masks outside the home (55.2%), preferring to walk by stairs then using the lift (61.1%). Our results are similar to a few other studies. ^17,20^

Male gender and divorced ones were the predictor of poor practice scores and age 51-60 years, private jobholders, and students were the predictors of the good practice scores. Our findings were similar to recently carried out investigations, which also mentioned the more responsible role of females as compared to males of the same age groups.^16, 11^ Our results are more or less similar to a Malaysian study which found good practicing among the general Malaysian population toward COVID-19.^8^ The practicing behaviour of old age people and students was better because initially, it was considered that it is the disease of the old and young people, and many of our study participants had the same misconceptions. Therefore, to avoid getting this infection, their practicing behavior was better.

Our results suggest that the majority of the study population was responding to this pandemic situation in a very responsible way; however, certain sections of society need more attentive education and mass awareness programs. As the invention of a vaccine against COVID-19 will take time, and so people will have to learn to live with COVID-19 in society. Recently, the World Health Organization (WHO) officials have announced that it is the probability that “this coronavirus may become just another endemic virus in our communities and this virus may never go away”. ^31^ No country can afford a ban on all commercial activities and closure of air routes and its borders for a more extended period. Therefore, we should continue working while observing WHO instructions of strict precautionary measures like personal hygiene, practicing good sneezing, and coughing etiquettes, frequently washing our hands with soap to protect ourselves and others. A clear policy should be constituted to deal with the psychological and mental wellbeing of the people in this pandemic. Our study results can be used by policymakers to set priorities in information campaigns on COVID-19 by the MOH/public health authorities and the mass media.

There are few limitations to our study, including its an online questionnaire-based study, so it is possible that respondents selected the most appropriate options, which is not being followed by them in real life. We used a convenience sampling technique, and in such studies, the respondents’ biases can’t be ignored. Besides, the study used an online questionnaire, so we could not reach the section of society that is not using the internet.

## CONCLUSION

Our study observed several predictors of misconceptions, knowledge, attitudes, and practices among the Saudi population. Our participants had a good realization of the impact of this pandemic.

## Data Availability

All data relevant to the study are included in the article.

## Acknowledgment

We are thankful to our medical students, Mahmood Abdullah Eid, Jinan Hikmat Msallati, Abdulelah Mugbil Hajer Alnafie, Nouf Khaleel Althagafi, Sultan Abdu Madkhali, Abdulrahman Omar Alzahrani for their help in collecting data.

## Ethical approval

This study was approved by the f the Unit of Biomedical Ethics of the King Abdulaziz University, Jeddah (Ref No. 187-20).

## Contributors

MB, TJ, Conceived and designed the study and wrote the manuscript SHA, AAM, ZJG, SHA contributed to acquisition of data, analysis of data, contributed to manuscript drafting TA,FB interpretation of the findings and manuscript editing. All authors revised and approved the final manuscript.

## Data availability

“All data relevant to the study are included in the article.”

---

**Table S1.** Study participants knowledge, attitude, and practice regarding COVID-19 pandemic.

| Statement | Frequency | Percentage |
| --- | --- | --- |
| **Knowledge** | **Correct Answer** |  |
| COVID-19 infection is caused by SARS-CoV-2. | 446 | 23.5% |
| COVID-19 infection is spread via respiratory droplets of the infected person. | 1641 | 86.5% |
| All community members are equally at risk for COVID-19. | 1111 | 58.6% |
| The best way of preventing spread of COVID-19 is social distancing | 1811 | 95.5% |
| The best way of preventing spread of COVID-19 is taking treatment | 1099 | 57.9% |
| Any type of group activity may spread this infection | 1785 | 94.1% |
| A symptomless COVID-19 patient (during incubation period) can't transmit infection | 1261 | 66.5% |
| The risk of getting infected when travelling by plane is higher | 1247 | 65.7% |
| This virus infection can be avoided by frequent hand washings by soap | 1705 | 89.9% |
| Advising Quarantine to passengers coming from infected areas is a good practice to avoid spread of infection | 1840 | 97.0% |
| Lockdown all over the country will control the spread of this virus | 1742 | 91.8% |
| Closing teaching institutions and shopping malls are effective ways of social distancing | 1825 | 96.2% |
| The most common cause of spread of this infection in any country is traveler from infected area | 1626 | 85.7% |
| Isolation period for infected people and those exposed to infection is 14 days | 1589 | 83.8% |
| **Attitude Questions** | **True Answer** |  |
| I am sure that COVID-19 infection will be overcome soon. | 1126 | 56.1% |
| We can overcome this problem by taking precautionary steps | 1878 | 93.6% |
| I understand that this infection is highly contagious | 1906 | 95.0% |
| It is my social responsibility to take safety measures in controlling spread of this infection. | 1957 | 97.6% |
| **Practice questions** | **Yes** |  |
| I am avoiding meeting my friends and relatives | 1784 | 89.9% |
| I am avoiding visiting crowded place | 1887 | 95.1% |
| I am avoiding using ATM machine. | 1028 | 51.8% |
| I prefer to walk by stairs then using lift | 1212 | 61.1% |
| I am using face mask outside the home | 1096 | 55.2% |
| I am using soap frequently for handwashing | 1806 | 91.0% |

**Table S2.** Study participants misconceptions and impact of COVID-19 pandemic.

| Statement (n=1985) | Correct answer |  |
| --- | --- | --- |
|  | n | % |
| An effective treatment is available against COVID-19. | 842 | 44.4% |
| This virus outbreak is only for short period. | 388 | 20.5% |
| COVID-19 be spread through frozen food | 606 | 31.9% |
| If a person's corona test is negative it means he is free from this virus | 318 | 16.8% |
| The outbreak of COVID-19 will be reduced during summer | 338 | 17.8% |
| Eating garlic can prevent corona virus infection | 828 | 43.6% |
| Females are more vulnerable to develop this infection | 1067 | 56.2% |
| Everyone should be tested for COVID-19 | 720 | 38.0% |
| Only older adults and younger people are at risk | 535 | 28.2% |
| Cats and dogs spread coronavirus | 916 | 48.3% |
| Disposable face masks protect against coronavirus | 621 | 32.7% |
| Hand dryers kill coronavirus | 856 | 45.1% |
| You have to be with someone for 10 minutes to catch the virus | 1018 | 53.7% |
| Rinsing the nose with saline protects against coronavirus | 899 | 47.4% |
| Thermal scanners can diagnose coronavirus | 645 | 34.0% |
| Sipping water every 15 minutes can protect corona infection | 825 | 43.5% |
| The coronavirus will die off when temperature rise in the spring | 551 | 29.0% |
| Coronavirus is the deadliest virus known to man | 920 | 48.5% |
| Flu and pneumonia vaccines protect against COVID-19 | 966 | 50.9% |
| **Impact questions** | **True** |  |
| I am very much scared of COVID-19 infection. | 925 | 46.3% |
| I am scared of food shortage during lockdown | 357 | 17.8% |
| COVID-19 is affecting my social, mental and psychological well beings. | 637 | 31.7% |
| The overall impact of this pandemic on me.<br>(participants were allowed to tick one or more options) |  |  |
| I have become careless | 106 | 5.3% |
| I have realized importance of life | 1784 | 86.2% |
| I have become more religious | 738 | 35.7% |
| The overall impact of this pandemic will be on: (participants were allowed to tick one or more options) |  |  |
| Healthcare system | 1417 | 68.5% |
| Economic condition of the country | 1583 | 76.5% |
| Economic condition of the people | 1284 | 63.8% |

## References

1. COVID-19 Dashboard by the Center for Systems Science and Engineering (CSSE) at John Hopkins University & Medicine. Available at: COVID-19 Dashboard by the Center for Systems Science and Engineering (CSSE) at John Hopkins University & Medicine. Available at: https://coronavirus.jhu.edu/map.html [Accessed on 21 May 2020]

2. Cucinotta D, Vanelli M. WHO Declares COVID-19 a Pandemic. Acta bio-medica 2020;91:157–160.

3. Mohamed RA, Aleanizy FS, Alqahtani FY, et al. Common co-morbidities are challenging in the diagnosis of Middle East Respiratory Syndrome (MERS-CoV) in Saudi Arabia. Science 2020;23:119–25.

4. Al-Tawfiq JA. Asymptomatic coronavirus infection: MERS-CoV and SARS-CoV-2 (COVID-19). Travel Med Infect Dis 2020;27:1–2.

5. Ebrahim SH, Ahmed QA, Gozzer E, et al. Covid-19 and community mitigation strategies in a pandemic. Br Med J 2020;368:m1066

6. Singh A, Purohit BM, Bhambal A, et al. Knowledge, attitudes, and practice regarding infection control measures among dental students in Central India. J Dent Educ. 2011;75:421–7.

7. Geldsetzer P. Use of rapid online surveys to assess People’s perceptions during infectious disease outbreaks: a cross-sectional survey on COVID-19. J Med Internet Res 2020;22:e18790.

8. Mohamad E, Azlan AA. COVID-19 and communication planning for health emergencies. Malaysian J Commun 2020;36.

9. Depoux A, Martin S, Karafillakis E, et al. The pandemic of social media panic travels faster than the COVID-19 outbreak. J Travel Med 2020;27:taaa031

10. World Health Organization. Coronavirus disease (COVID-19) advice for the public: Myth busters. January 2020. Available at: www.who.int/emergencies/diseases/novel-coronavirus-2019/advice-for-public/myth-busters. [Accessed on 25 March 2020].

11. Zhong BL, Luo W, Li HM, et al. Knowledge, attitudes, and practices towards COVID-19 among Chinese residents during the rapid rise period of the COVID-19 outbreak: a quick online cross-sectional survey. Int J Biol Sci 2020;16:1745–52

12. Faasse K, Newby JM. Public perceptions of COVID-19 in Australia: perceived risk, knowledge, health-protective behaviours, and vaccine intentions. MedRxiv. 2020

13. Ortiz-Ospina E. The rise of social media; 2020. Available from: https://ourworldindata.org/rise-of-social-media. [Accessed on 12 May 2020].

14. Vraga EK, Bode L. Defining misinformation and understanding its bounded nature: Using expertise and evidence for describing misinformation. Political Commun. 2020;37:136–44.

15. Vosoughi S, Roy D, Aral S. The spread of true and false news online. Science 2018;359:1146–51.

16. Naser AY, Dahmash EZ, Alwafi H, Alsairafi ZK, Al Rajeh AM, Alhartani YJ, Turkistani FM, Alyami HS. Knowledge and practices towards COVID-19 during its outbreak: a multinational cross-sectional study. MedRxiv. 2020.

17. Geldsetzer P. Knowledge and perceptions of coronavirus disease 2019 among the general public in the United States and the United Kingdom. Ann Intern Med. 2020.

18. Tenkorang EY. Myths and misconceptions about HIV transmission in Ghana: what are the drivers?. Cult Health Sex 2013;15:296–310.

19. Ahmad T, Haroon, Baig M, et al. Coronavirus Disease 2019 (COVID-19) Pandemic and economic impact. Pak J Med Sci 2020;36(COVID19-S4):S1-S6.

20. Haleem A, Javaid M, Vaishya R. Effects of COVID 19 pandemic in daily life. Curr Med Res Prac 2020.

21. Gros C, Valenti R, Valenti K, et al. Strategies for controlling the medical and socioeconomic costs of the Corona pandemic. arXiv:2004.00493. 2020.

22. Chew QH, Wei KC, Vasoo S, et al. Narrative synthesis of psychological and coping responses towards emerging infectious disease outbreaks in the general population: practical considerations for the COVID-19 pandemic. Singap Med J. 2020.

23. Holmes EA, O’Connor RC, Perry VH, et al. Multidisciplinary research priorities for the COVID-19 pandemic: a call for action for mental health science. Lancet Psychiat 2020;7:547–560.

24. Kobayashi T, Jung S-M, Linton NM, et al. Communicating the Risk of Death from Novel Coronavirus Disease (COVID-19). J Clin Med 2020;9:580.

25. Newby JM, Haskelberg H, Hobbs MJ, et al. The effectiveness of internet-delivered cognitive behavioural therapy for health anxiety in routine care. J Affect Disord 2020;264:535–42.

26. Giao H, Nguyen TNH, Tran VK, et al. Knowledge and attitude toward COVID-19 among healthcare workers at District 2 Hospital, Ho Chi Minh City. Asian Pac J Trop Med 2020;13.

27. Shi Y, Wang J, Yang Y, et al. Knowledge and attitudes of medical staff in Chinese psychiatric hospitals regarding COVID-19. Brain Behav Immun Health. 2020: 100064.

28. Hussain ZA, Hussain SA, Hussain FA. Medical students’ knowledge, perceptions, and behavioral intentions towards the H1N1 influenza, swine flu, in Pakistan: a brief report. Am J Infect Control 2012;40: e11–e13.

29. Clements JM. Knowledge and behaviors toward COVID-19 among US residents during the early days of the pandemic. MedRxiv. 2020.

30. Pennycook G, McPhetres J, Bago B, et al. Predictors of attitudes and misperceptions about COVID-19 in Canada, the UK, and the USA. PsyArXiv. April 2020. doi:10.31234/osf.io/zhjkp.

31. BBC NEWS. Coronavirus may never go away, World Health Organization warns. Avaialbe at: https://www.bbc.com/news/world-52643682. [Accessed on 15 May 2020]

